# Exploring the relationship between food environment and preferences among school children in a low socio-economic community in Sri Lanka: A GIS based assessment

**DOI:** 10.1101/2023.09.10.23295336

**Authors:** Chamil Senevirathne, Prasad Katulanda, Padmal de Silva, Dilini Prashadika, Lalith Senarathne

**Affiliations:** Department of Health Promotion, Faculty of Applied Sciences, Rajarata University of Sri Lanka; Department of Clinical Medicine, Faculty of Medicine, University of Colombo; Department of Statistics, National Institute of Health Sciences, Kaluthara, Sri Lanka; Department of Computing Sciences, Faculty of Applied Sciences, Rajarata University of Sri Lanka

## Abstract

Food environment in school neighborhood plays a crucial role in manipulating food choices of school children. This study investigated the relationship between food environment in neighborhood and dietary practices of government school students in a low socio-economic setting, Sri Lanka. This cross-sectional study surveyed the neighborhood food environment of selected schools (n=30) in Monaragala District, Sri Lanka, using Geographical Information System (GIS) data, and collected dietary information from a representative sample of school children (n=603). Chi-square and spearman correlation tests were performed using SPSS version 23.0 to estimate the associations between food environment and BMI of students, while ArcGIS 10.4.1 was used to analyze GIS data of the study.

Majority of students (35.5%) were 15 years old and approximately 51% were females. Mean BMI of study participants was 18.14 (± 3.28). More than 90% of outlets within close proximity sold unhealthy foods. Consumption of confectionaries was 72.3% of students whereas healthy foods choices was ranged from 5% to 12%. A positive correlation between consuming unhealthy food and distance to the outlets from the school was observed (p<0.05). Risk of consuming low nutrition food found to be doubled (OR = 2.47, 95% CI: 1.52-3.89) among students studied in schools where larger proportion of energy dense food sold in closer proximity. In conclusion, density and the proximity of the outlets that sold food in low nutrients in school neighborhood environment were positively associated with choosing unhealthy food items by students.

## Introduction

Neighborhood food environment in school contains the physical infrastructure and overall surroundings within adjacent to school premises where food is both sold and consumed. This encompasses the spectrum of food items available in these locations, considering not only their availability but also the nutritional composition they offer [1]. The presence of unhealthy food options in close proximity to schools significantly shapes the dietary choices of school children, particularly during their adolescence. This phase of life pivotal for the development of both cognitive and physiological aspects that impact an individual’s overall well-being across the lifespan [2–3]. Positive lifestyle and behavior practices during this period reduce the risk of chronic diseases in early adulthood [4]. Marketing strategies of the food industry influence school children to reach food destinations such as fast food huts, grocery outlets and cafeteria to fulfill their day to day dietary requirements [5]. A positive association between availability of food outlets in closer proximity and eating habits, nutritional status of people across different settings and populations is already established [6–7]. A recent study demonstrated that neighborhood food destinations fulfill significant amount of daily caloric intake of school children in USA [8]. However, presence of energy dense foods which are promoted through child focused marketing, substantially influence food choices of school children [9]. Several studies have shown that children and adolescents who have easy access to unhealthy food outlets near their schools tend to have a higher consumption of fast food, sugary beverages and snacks. This is often associated with an increased risk of obesity and related health issues [10–13]. The presence of healthy food on the other hand, has been linked to better dietary choices as well. Therefore, school neighborhood food environment plays a crucial role in influencing on eating pattern, food preference and further school children as they are exposed to wide range of unhealthy food and beverage items during the school hours.

Sri Lanka is a middle income country with diverse socio-economic background across different communities. Approximately 20% of the population represents school age children and adolescents [14]. Majority (57.3%)of school children are provided free education through 10,165 government schools, while others attend private (37.2%) aided (8.5%) and other schools (3.3%) [15], and all the provincial schools are categorized in to four types namely AB, 1C, type 2 and type 3 based on the infrastructure facilities. Sri Lankan government introduced a school canteen policy in 2006 to ensure the access to nutritional food within school premises. This policy is mainly focused on changing the food environment within school premises. Although, having a policy to regulate the food availability in the school premises is a vital part in improving nutritional practices of children, school neighborhood should also be taken in to consideration when changing the school food environment. Comprehending the food environment, encompassing the nutritional value of accessible food items both within and their surrounding neighborhoods, holds significant sway over the nutrition wellbeing of school children. While handful of studies have probed in to the school nutritional environment and its correlated factors in urban settings in Sri Lanka [16], there remains a notable dearth of evidence concerning economically disadvantaged communities. Consequently, there exists discernible gap in the body of evidence pertaining to the research on school food environment in Sri Lanka. Addressing this gap holds paramount importance in generating the essential additional evidence required for the efficacious implementation of food environment policies aimed at schools. The objective of this study was to explore the association between neighborhood food environment and dietary practices of school children within a low-socioeconomic community in Sri Lanka. This exploration was conducted through the lens of geographical accessibility, utilizing Geographic Information System (GIS) as a fundamental tool.

## Methodology

### Study Design

A cross-sectional study was conducted in government schools under the provincial department of education in Moneragala Education zones of Moneragala District of Sri Lanka. Poverty indicators in Sri Lanka [17] was used to define economic status of the study.

### Study Population

#### Primary target population

School children from grade eight to eleven (age from 13 to 16 years) of government schools in Moneragala education division in the Uva province of Sri Lanka were selected as study population. Students with cognitive disability and living with chronic diseases requiring long term drug treatments and follow-up were excluded from the study.

#### Secondary target population

outlets which sold ready to eat food items, process food, confectionaries and other type of snacks were considered for the data collection. All the outlets that sold food items (except mobile carts and mobile food sellers), located within 300 meters from the school border were selected for the assessment. Other outlets that did not sell any type of food were excluded from the survey.

### Sample size calculation

In low socio-economic groups, proportion of students who access neighborhood food outlets were considered for the sample size calculation. Since there were no previous studies to report the prevalence, sample size was calculated to detect at least 50% prevalence within the population. Sample size was calculated using the formula for cross sectional studies from N= z^2^p (1-p)/d^2^ [18]. Together with the hypothesized prevalence of accessing neighborhood food outlets among school children (50%), 0.05 level of significance (precision d = 5%), Z= 1.96 standard normal deviation at 95% CI was included for the calculation.

Previous studies suggested including the designed effect 1.5 to ensure the statistical validity of the sample selection procedure. In order to minimize the error due to clustering, the calculated sample size was multiplied by the design effect (D) which was taken as 1. 516 [19].Then the sample size was 576 and to compensate for the non-response rate of 10% another 58 were added to the sample, and Final sample size was 630. Multistage stratified cluster sampling was used to select the required number of clusters (n=30). Describing selecting the secondary sample, all the outlets that sold food, located within 300 meters’ radius from the school main gate was considered for the data collection.

### Data collection procedure and Study instruments

The validated version of Global School Health Survey (GSHS) questionnaire was used to collect demographic and dietary information of the participants [20]. This interviewer-administered questionnaire collected two types of information, the first part investigated socio-demographic data including age, gender, parent’s education and family income while second part collected information in relation to nutrition related practices such as consumption of deep-fried food, confectionaries, sweets, fruits and vegetables, etc. in the previous 30 days. The questionnaire was translated to Sinhala and Tamil languages and then back-translated to English to ensure the accuracy of the content. The same one was pre-tested with a sample in order to observe understandability and accuracy of responses.

An observation check list which was developed based on the recommendations of the school canteen policy [21] was utilized to determine the availability of different food types in school proximity. Each shop was visited by the investigator to identify the availability of healthy and unhealthy food, and recorded under selected category of the check list. Healthy and unhealthy food was defined according to the recommendations of the school canteen policy [21]. Food environment was defined as the all the outlets selling food within 300 meters from the school. A sample geographical analysis was done by the investigator prior to determine the buffer distance. Since the majority of students resided in walking distance from their schools, maximum radius for the data collection was decided as 300 meters. In addition to determining food context of the outlets, coordinates of the outlets and schools captured by using Garmin ETrex 10 GPS device and then tabulated in Microsoft Excel 2010.

In order to establish physical measurements of study participants, height and weight were measured to the nearest 0.1 cm and 0.1 kg by trained data collectors. Standardized procedures for anthropometric measurements were used to obtain both measurements [22].

### Data Analysis

During the analysis phase, descriptive methods were employed in alignment with the study objective. The examination of relationships between chosen variables and dietary preference of the study participants was conducted through the Chi-square & Spearman Correlation tests. These statistical tests were utilized to ascertain the degree of association between the variables and the dietary choices of the participants. Statistical significance was assessed at (95%, CI) p < 0.05 level. Statistical Package on Social Studies (SPSS 23.0) was used to analyze the data.

In order to analyze GIS data, the present study used ArcGIS 10.4.1 software to analyze spatial neighborhood information. GPS coordinates of outlets and schools were used to create point layer on ArcMap and using these two layers could be analyzed distance from each school to outlets. Based on every school location created multiple buffer zones and these buffer zones clearly show surrounding food outlets of the school environment. Shop density and proximity were defined as the operational variables for the number of outlets located within given buffer zones and distance from the school respectively.

### Ethical Clearance

The study made necessary measures to ensure ethical validity of the study. All the participants were provided an information sheet about the study procedure, and obtained the written consent after providing opportunity to ask questions. In addition, ascent consent was taken from parents and guardians of school children Steps were taken to maintain the confidentiality of data and privacy of the study participants. Study protocol was reviewed under and approved by the Ethical Review Committee of Faculty of Medicine, University of Colombo; **Approval number EC-14-105**.

## Results

With response rate of 95.7%, the study recruited a cohort of 603 students, selected from 30 schools to ensure representation across all three education zones within Moneragala District in Sri Lanka. Among the selected sample, a predominant proportion (35.5%) comprised 15 years old, while approximately 51% were females. Notably, more than half (56.4%) of the enrolled students hailed from low-income families as evidence by data presented in table 1.

**Table 1:** Socio-demographic characteristics of participating students.

| Characteristic | Frequency | % |
| --- | --- | --- |
| Gender |  |  |
| • Female | 311 | 51.6 |
| Age group |  |  |
| • 13 years | 95 | 15.8 |
| • 14 years | 188 | 32.2 |
| • 15 years | 214 | 35.5 |
| • 16 years | 106 | 17.5 |
| Mother's education level |  |  |
| • No education | 112 | 18.6 |
| • Primary education | 246 | 40.8 |
| • Secondary education | 245 | 40.6 |
| Father's education level |  |  |
| • No education | 145 | 24.0 |
| • Primary education | 289 | 48.0 |
| • Secondary education | 169 | 28.0 |
| Monthly income |  |  |
| • Low | 340 | 56.4 |
| • Middle | 160 | 26.5 |
| • High | 103 | 17.1 |
| Total | 603 | 100.0 |

As presented in table 2, the mean BMI for males were 17.6 Kgm^-2^ (± 3.4) while for females it was 18.5 Kgm^-2^ (± 3.1). Remarkably, a substantial majority of the study participants (65.7%) were classified as underweight, in contrast to a modest 8.3% who fell into the overweight category. The prevalence of underweight conditions was notably elevated among male students (74.7%), indicating a significant disparity (p <0.001) in comparison to their female counterpart (57.1%).

**Table 2:** Distribution of students according to their weigh status.

|  | Nutritional status |  |  |  |  |  |  |  |  |
| --- | --- | --- | --- | --- | --- | --- | --- | --- | --- |
|  | Underweight | % | Normal weight | % | Over weight | % | Total | % |  |
| Male | 219 | 74.7 | 56 | 19.1 | 18 | 5.2 | 293 | 100 | df=2<br>X<sup>2</sup>=19.9<br>P=.001 |
| Female | 177 | 57.1 | 101 | 32.5 | 32 | 10.4 | 310 | 100 |  |
| Total | 396 | 65.7 | 157 | 26.0 | 50 | 8.3 | 603 | 100 |  |

The table 3 displays the distribution of food outlets across the individual buffer zones. The analysis incorporated a total of 97 outlets situated within 300 meters’ radius. It is notable that a significant proportion of these outlets were concentrated within 100 meters’ buffer zone, with comparatively lower count observed within broader 300 meters’ buffer zone. Further, Fig 1 visualize the physical characteristics of the school neighborhood food environment.

**Figure 1:**
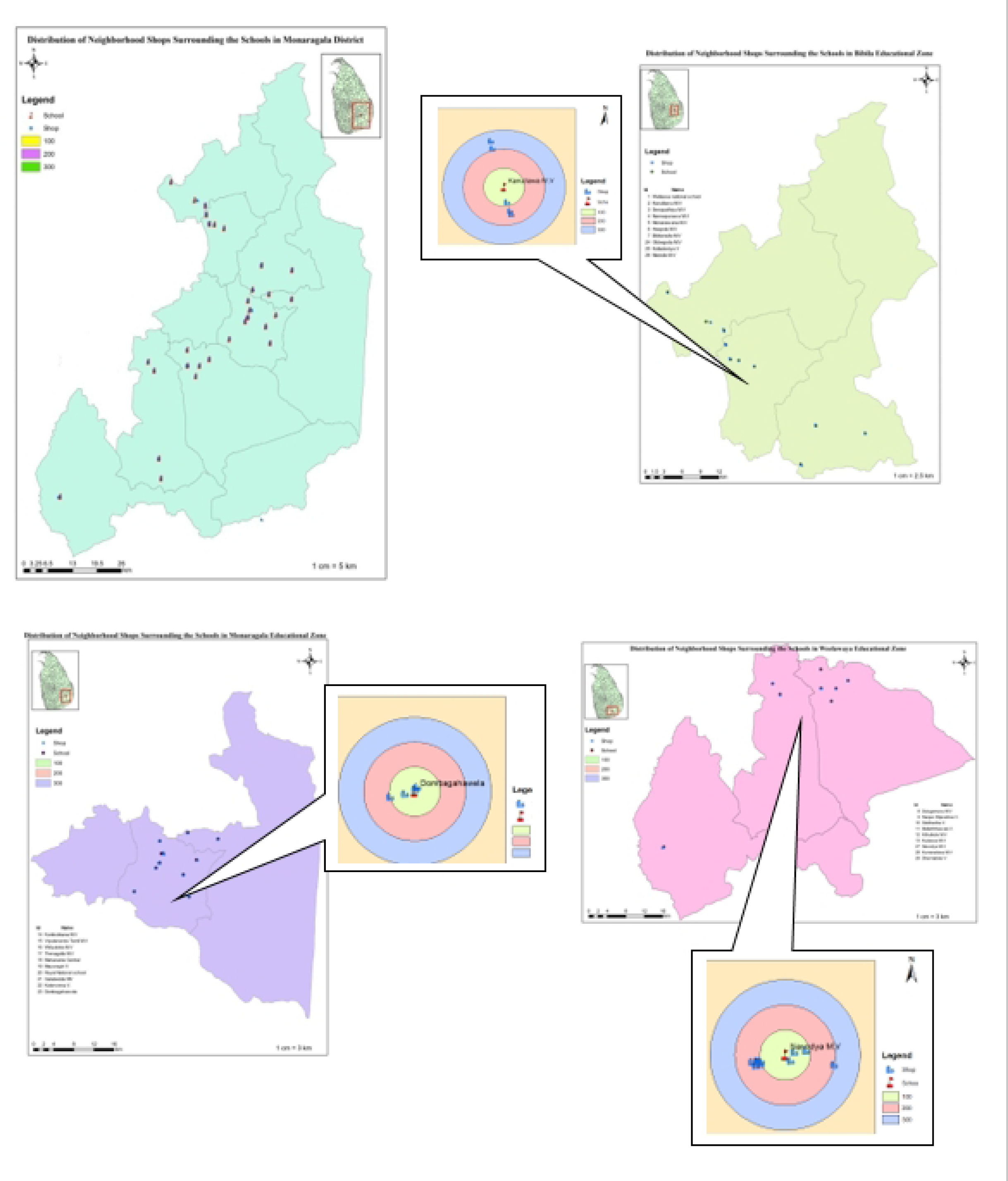
Distribution of shops and schools in three education zones.

**Table 3:**
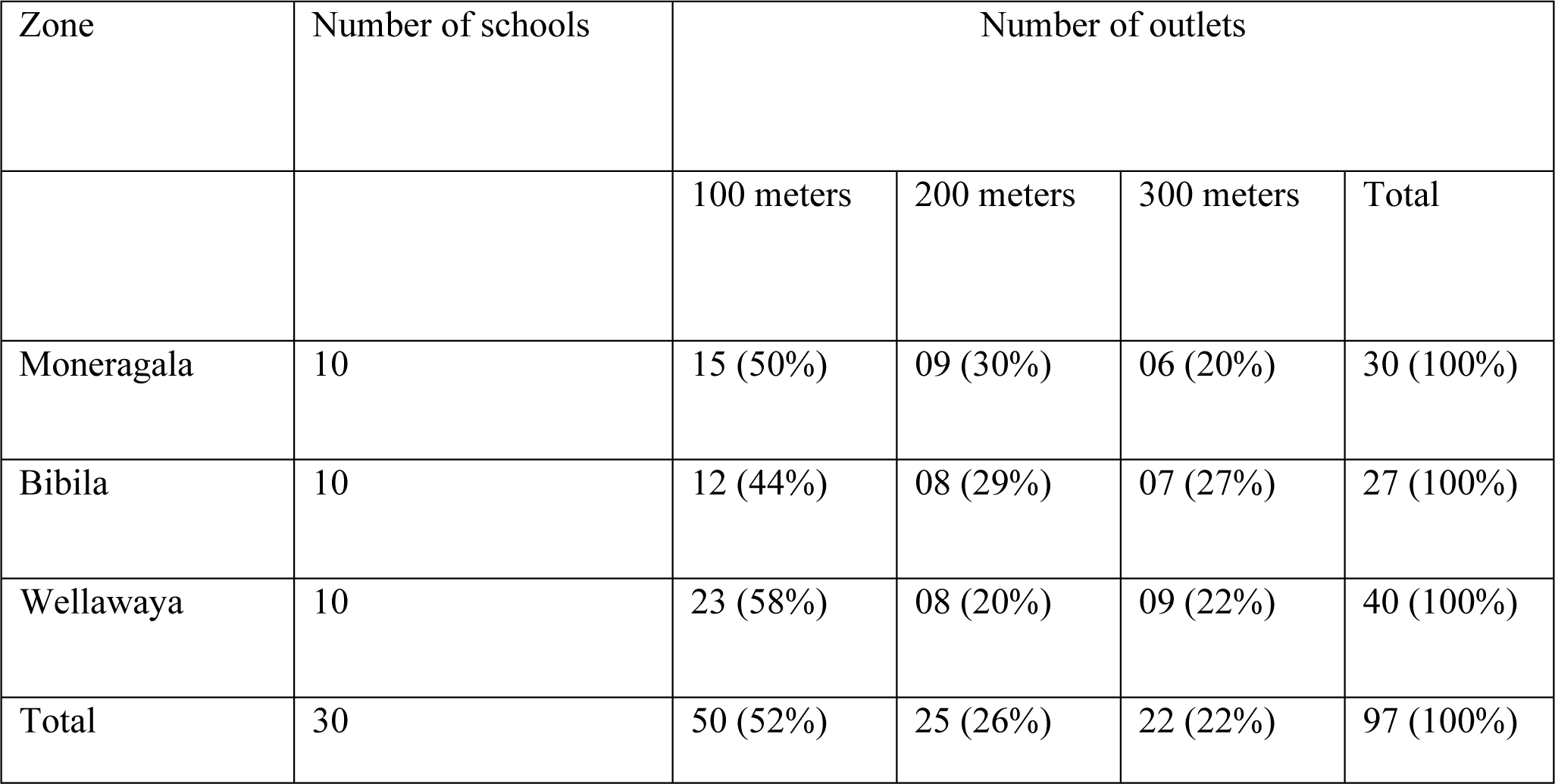
Shop distribution in given buffer zones in each educational zone.

| Zone | Number of schools | Number of outlets |  |  |  |
| --- | --- | --- | --- | --- | --- |
|  |  | 100 meters | 200 meters | 300 meters | Total |
| Moneragala | 10 | 15 (50%) | 09 (30%) | 06 (20%) | 30 (100%) |
| Bibila | 10 | 12 (44%) | 08 (29%) | 07 (27%) | 27 (100%) |
| Wellawaya | 10 | 23 (58%) | 08 (20%) | 09 (22%) | 40 (100%) |
| Total | 30 | 50 (52%) | 25 (26%) | 22 (22%) | 97 (100%) |

Over 90% of the outlets located in the immediate vicinity of the school included in the study were observed to primarily offer ready to eat deep fried snacks (such as bites, short-eats), processed food items, bakery products such as bread and buns, as well as sugary treats such as sweetmeats, toffee and chocolates. Surprisingly a mere 12 out of total 97 outlets surveyed (12.8%) were found to have fruits available for sale. Interestingly none of the assessed food outlets offered any healthy beverage options such as fresh milk or freshly squeezed fruit juice (Fig 2).

**Figure 2:**
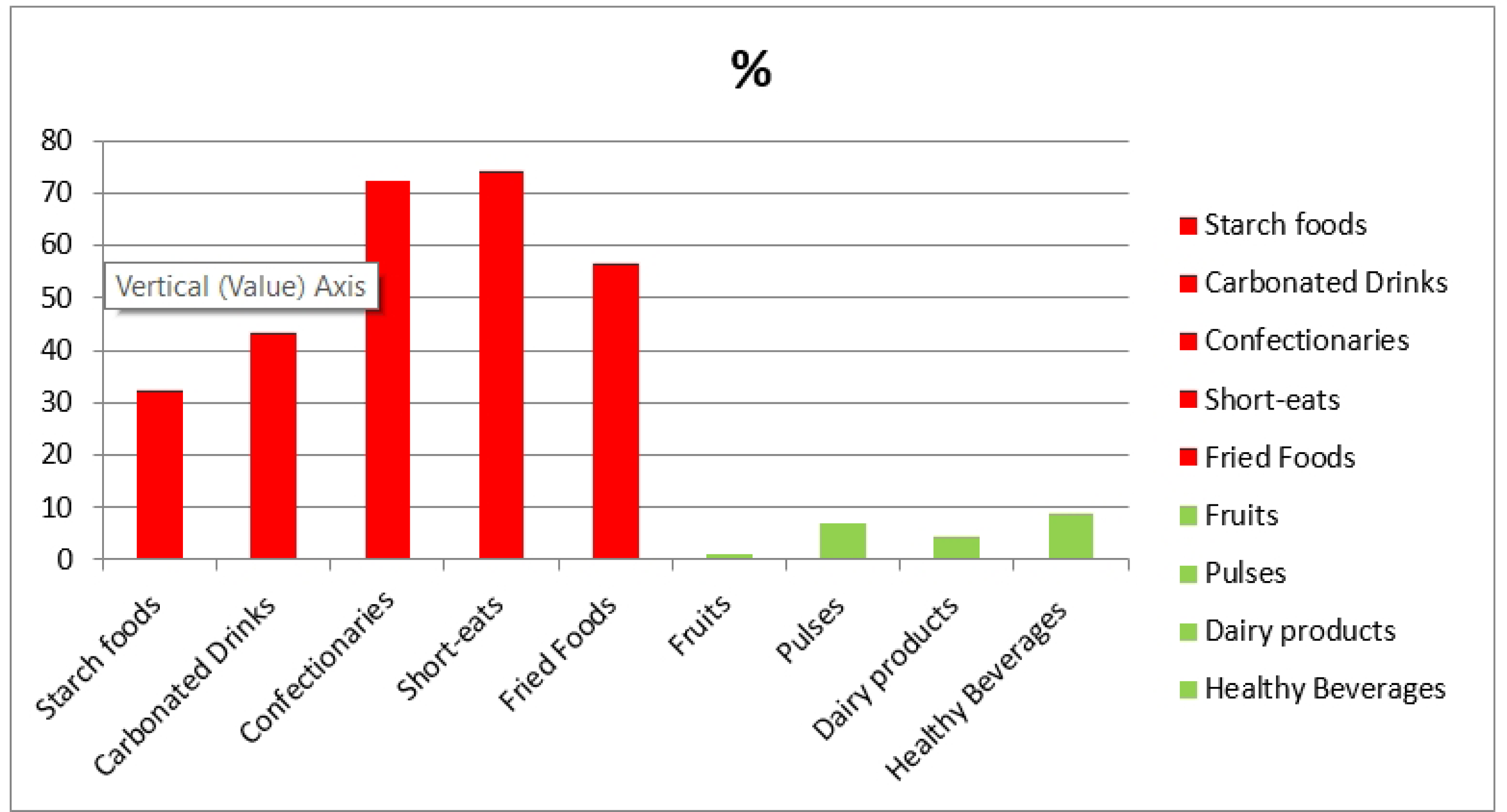
Different types of snacks consumed by the study participants.

This study indicated that approximately two third (58.1%) students consumed snacks from food outlets in the school premises. Notably, there was a discernible gender-based distinction, with a higher prevalence of food consumption from these outlets observed among male students compared to their female counterpart. The findings also indicated a noteworthy dietary trend, with carbonated drink consumption (38.5%) significantly outweighing the intake of healthier beverage alternatives (8.8%), showcasing nearly fourfold difference. Furthermore, an overwhelming 72.3% of students reported the consumption of confectionaries, while 37.5% reported their consumption of starch-rich food (Table 4).

**Table 4:** Classification of food outlets offerings based on availability of diverse food items.

| Food items | Number of outlets | % |
| --- | --- | --- |
| ***Unhealthy food*** |  |  |
| Deep fried food | 90 | 92.8 |
| Short-eats | 90 | 92.8 |
| Confectionaries | 97 | 100 |
| Bakery products | 97 | 98.9 |
| Carbonated beverages | 80 | 82.5 |
| ***Healthy food*** |  |  |
| Pulses | 00 | 00 |
| Fruits | 12 | 12.3 |
| Dairy products (yoghurt/curd) | 68 | 70.1 |
| Healthy beverages (Herbal drinks/fresh milk/fruit juice) | 00 | 00 |

When considering food outlets with unhealthy food available within 100 meters’ radius, the estimated odds of consuming unhealthy food among school students significantly increased to 2.47 (OR = 2.47, 95% CI: 1.52-3.89), compared to schools situated amidst fewer outlets energy dense food. Similarly, the presence of an increased number of outlets selling unhealthy food within a 200 meters’ buffer zone doubled the likelihood of selecting unhealthy food items. (OR=1.99 95% CI 1.44-2.93) when compared to schools with a lesser availability of outlets with unhealthy food (Table 5).

**Table 5:** The correlation between the presence of unhealthy food within the school local surrounding and the dietary habits of school children.

| Availability of unhealthy food | Consuming energy dense food from outlets Yes vs No<br>OR (95% CI) | p |
| --- | --- | --- |
| Within 100 meters (Yes vs No) | 2.47 (1.52-3.89)*** | 0.000 |
| Within 200 meters | 1.99 (0.46-2.9)** | 0.036 |
| Within 100 meters | 1.63 (0.5- 4.75) | 0.092 |

As presented in table 6. a statistically significant correlation has been established between consumption of unhealthy food among school children and the average distance separating the outlets from the respective schools within each buffer zone. Specifically, the analysis indicates a positive relationship between the prevalence of unhealthy food consumption among the study participants and the proximity of outlets located within 100 meters’ and 200 meters’ buffer zones.

**Table 6 :** correlation between consumption of unhealthy food among study participants and proximity of outlets.

| Density and proximity | (Coefficient 95% CI) | P |
| --- | --- | --- |
| Number of outlets within 100 meters boundary | 0.344* | 0.031 |
| Number of outlets within 200 meters boundary | 0.358* | 0.033 |
| Number of outlets within 300 meters boundary | 0.032 | 0.292 |
| Distance to the nearest shop from the school | -0.312* | 0.034 |

## Discussion

Our study revealed that nearly all food outlets situated within a 300-meter proximity were engaged in selling unhealthy food, whereas only a limited number of outlets adhered to the healthy item criteria defined by the school canteen policy in Sri Lanka. On the other hand, this study observed that larger proportion of children (58.1%) consumed food from outlets located in school neighborhood environment, and discretionary snacks, and most types of food consumed by the were unhealthy. Moreover, this study indicates that the food environment of school settings has a significant relationship with the dietary choices of children in those schools. This relationship is reinforced as the density and proximity of food outlets in the school neighborhood increased.

In this study, higher percentage of unhealthy food availability in the school proximity was observed. These findings are in line with the previous studies that reported a higher availability of unhealthy food items in the school environement [23–24]. In agreement with existing literature [25–26] our study also found males were more in to consuming food from the nearest food outlets. Further, sugary beverages, bakery items and deep-fried foods were the most popular food items whereas less demand for healthy food options were observed among school children. Although present study did not explore the reasons for higher availability of unhealthy food in the school food environment, a recent study further suggested that students may be culturally influenced for consuming such energy dense foods [27]. On the other hand, recent studies had demonstrated that higher fast food consumption among school children may be due to the availability of such food items in the school environment [26–28]. Hence, it is possible to argue that increasing availability of unhealthy food increases the demand for such food by school children. Our study re-affirmed this fact as higher consumption of energy dense food from the closer proximity was observed among participants.

This study uncovered a significant correlation between opting for unhealthy choices and the proximity of unhealthy food options within a 100 meters’ radius from the school premises. This association is in line with the findings from previous studies conducted in the United Kingdom [29] and South Korea [30] These findings underscore the powerful influence of the local food environment ‘s density and immediate availability on the dietary preference exhibited by students. Such insight holds the potential inform targeted interventions aimed at promoting their healthier food options within close proximity to schools. Although, the present study involved school children from low and middle income country, findings are comparable with the studies from high income countries [31]. Therefore, this study re-emphasizes the need of considering evidence from settings with varied socio-economic status in developing polices on the impact of school food environment.

Existing evidence claim that solidity of food outlets coupled with availability of unhealthy food options influences dietary choices of school children [32]. In contrast, this study found that availability of unhealthy foods within 100 meters and 200 meters buffer zones doubled the risk of consuming energy dense food by school children. This finding is in agreement with a previous study that established an interaction between the availability of unhealthy food in walk shed food outlets and the dietary behaviour of school going adolescents [33]. In consistent with recent findings, this study indicates that outlets density in first and second buffer zones was positively associated with the food consumption of the students [31–33]. Hence, present study re-emphasizes that proximity can be attributed to increase the unhealthy food choices of the school children. However, several studies have found no relationship between school environment and the consuming energy dense food by school children [34–36]. This disparity may be due to the factor that most of the studies being compared, had framed to urban communties in developed countries, and assessed mostly the super-market, convinient stores as well as fast food restuerents where students may not have access for purchasing food. On the other hand, this study examined only the food destinatoins located within walking distance, whereas most of the studies from highincome settings considered comparatively extended buffer zones for the analysis based on available commuting facilities. The findings of our study demonstrated the importance of enhancing current nutrition related policies through the incoorporation of regional factors which encompass demographic, socio-economic and cultural factors as well as individual characteristics. This approach ensures equatable distribution of policy benefits across the entire country.

In this study, substantial proportion of study participants were found to be underweight, and it can be contented that consumpton of energy-dense food is significance in attaining their ideal weight. Nevertheless, empirical evidence demonstrates that consumption nutrients-deficient food serves as pivotal factor in the nutritional shift from underweight to overweight or obese[37]. In addition to that, recent evidence affirm that exposure to junk food options during the childhood severly influence on their dietary cuture, food preference and attitudes towards unhealthy dietary practices [38]. Further, frequent fast food consumers likely to decrease odds of sufficient dietary habits such as lower intake of fruits and vegitables [38–39]. Our study confirms this reality by highlighting a reduced inclination towards healthier food options ad advised by the school canteen policy in Sri Lank .

The key strength of our study lies in its pioneering use of Geographical Information System (GIS) to offer a comprehensive evaluation of how the school neighborhood food environment influences dietary preference of school children in rural Sri Lanka. In addition to that this study introduced a mechanism to define, characterize and quantify the school food environment through the application of Geographical Information System. Moreover, the study investigated the influence of proximity and density of food outlets on nutritional practices of school children in low income community. Therefore, our findings address the knowledge gap of the impact of school food environment on eating habits of school children in low income communities in Sri Lanka

One of the limitations is that this study examined all food outlets within the defined buffer zone, in contrast to many other studies that classified these food selling establishments based on criteria such as range of food choices, geographical placement and consumer oriented amenities [40–41]. Consequently, this approach potentially constrains our ability to fully explore how the food preference of school children are influenced by physical attributes of these food outlets. Present study also did not consider factors such as purchasing power of students, affordability of food, quantity of consuming snacks and other motivation of consuming food from the neighborhood outlets. Another limitation of the study pertains to the method used in defining the food preference of school children, which relied on assessing the frequency of consumption of unhealthy food items without considering quantitative analysis of food consumption pattern. Therefore, future studies are encouraged to employ methods such as 24 hours’ dietary recall and food frequency questionnaire that could provide more detailed understanding of the dietary habits and preference of the study population.

## Conclusion

The present study delved into the dynamics of food availability within defined buffer zone surrounding the school environment, examining the variety of food options presented in the immediate vicinity. Furthermore, it aimed to establish connections between the density and proximity of food outlets and the dietary choices made by students. The results emphasized a concerning trend: a pronounced prevalence of unhealthy food choices within the food outlets adjacent to schools. Strikingly, more than half of the surveyed population opted to obtain their food from these nearby school neighborhood food outlets. The study findings also revealed a notable preference for energy dense food items among the study participants. In essence, this study emphasized the substantial influence wielded by the characteristics of the neighborhood food environment, specifically their density and proximity.

### Recommendations

Findings of the present study highlight that healthy food environment around the school premises is important to ensure the nutritional behaviour of school children in low income community. Therefore, measures to create and sustain healthy food environment should be undertaken at school level as well as policy level. school canteen policy which is the only available policy for shcools covers merely the food environment within school premises in Sri Lanka. However, it does not address the influence of crucial neiborhood food environment as highlighted by this study. Therfore it is recommended to expand exisiting policies regarding school food environment beyond school premises to include school neiborhood. Such initiatives can be based on the exisiting similar policies to address other public health issues related to adolesccents; National Alcohol and Tobacco Act (NATA) [42] which prevent selling cigerette withing the premises and close porximity of schools. In addition to establishing policies and mechanism to monitor policy, encouraging health promotion initiatives for behaviroual changes among students my contribute to address issues revealed in this study.

## Data Availability

Data is securely maintained with stringent measures in safeguard its confidentiality. Nevertheless, the author is ready available to provide the data upon request any time.

## Acknowldegement

The authors of the study express their heatfelt gratitude to all participants of the study, including school children and shopkeepers, whose enthusiastic participation was invaluable. Special recognition is extended to school staff for generously granting permission to access the school premises for the purpose of data collection. Lastly, our sincere appreciation goes to the administrative body of the Department of Educaiton, Uva Province Sri Lanka, for their kind authorization, enabling the study to be conducted across three division.

## References

1. Food and Agriculture Organization of the United Nation . “Healthy food environment and school food”. United Nation. Available from: https://www.fao.org/school-food/areas-work/food-environment/en/#:∼:text=The%20school%20food%20environment%20refers,the%20nutritional%20content%20of%20these. (Accessed 20/03/2022).

2. Fox MK, Dodd AH, Wilson A, Gleason P. Association between School Food Environment and Practices and Body Mass Index of US Public School Children. American Dietetic Association. 2009; 109.

3. Chaudhary A, Sudzina F, Mikkelsen BE. Promoting Healthy Eating among Young People—A Review of the Evidence of the Impact of School-Based Interventions. Nutrients. 2020; 12.

4. Lytle LA, Kubik MY. Nutritional issues for adolescents. Best Pract. Res. Clin. Endocrinol. Metab. 2009; 17: 177–189.

5. Buck C, Bornhorst C, Pohlabeln H, Huybrechts I, Pala V, Reisch L, Pigeot I. Clustering of unhealthy food around German schools and its influence on dietary behavior in school children: a pilot study. International Journal of Behavioral Nutrition and Physical Activity. 2013;10.

6. Caspi CE, Sorensen G, Subramanian SV, Kawachi I. The local food environment and diet: a systematic review. Health Place. 2012; 18: 1172–1187.

7. Feng J, Glass TA, Curriero FC, Stewart WF, Schwartz BS. The built environment and obesity: a systematic review of the epidemiologic evidence. Health Place. 2010; 16: 175–190.

8. Jia P, Xue H, Cheng X. et al. Effects of school neighborhood food environments on childhood obesity at multiple scales: a longitudinal kindergarten cohort study in the USA. BMC Med. 2019; 17: 99.

9. Harris J, Bargh JA, Brownell D. Priming Effects of Television Food Advertising on Eating Behavior. Health Psychology. 2009; 28: 404–413.

10. Sanchez BN, Vaznaugh EV, Uscilka A, Baek J, Zhang L. Differential Associations Between the Food Environment Near Schools and Childhood Overweight Across Race/Ethnicity, Gender, and Grade. American Journal of Epidemiology. 2012; 175: 1284–1293.

11. Nollen N, Befort C, Snow P, Daly CM, Ellerbeck F, Alhuwalia J. The school food environment and adolescent obesity: Qualitative insight from highschool principals and food service personals. International Journal of Behavioral Nutrition and Physical Activit. 2007; 4: 279–281.

12. An R, Sturm R. School and Residential Neighborhood Food Environment and Dietary Intake among California Children and Adolescents. American Journal of Preventive Medicine. 2012; 42:129–135.

13. Williams J, Scarborough, P, Matthews A, Cowburn G, Roberts N, Rayner M. A systematic review of the influence of the retail food environment around schools on obesity-related outcomes. Obesity reviews. 2008; 15: 359–374.

14. Sri Lanka Demographic and Health Survey, Department of Senses and Statistics, Ministry of National Policies and Economic Affairs. (Accessed 22/03/2022). http://www.statistics.gov.lk/Resource/en/Health/DemographicAndHealthSurveyReport-2016-Contents.pdf.2016.

15. Ministry of Education, Sri Lanka (MoE). Sri Lanka Education information, Ministry of Education [Online]. Nutrition Coordination Division. (Accessed 17/02/2022). http://www.moe.gov.lk/web/index.php?option=com_content&view=article&id=300&Itemid=114&lang=env2012.

16. Sirasa F, Mitchell L, Harris N. Dietary diversity and food intake of urban preschool children in North-Western Sri Lanka. Maternal & child nutrition. 2020; 16(4): e13006.

18. Department of Census and Statistics (DSC). Poverty Indicators-2019. Ministry of Economic Policies and Plan Implementation. April 2022(online). PovertyIndicators-2019.pdf (statistics.gov.lk). [Accessed 22/03/2022].

18. Lawanga SK, Lemeshow S. Sample size determination in health studies : a practical manual. In: World Health Organization (ed.). 1991. Genevia World Health Organization.

19. Gabler S, Hader S, Lynn P. Design Effects for Multiple Design Samples In: Essex, U. O. (ed.). United Kingdom Institute for Social and Economic Research. 2005.

20. Ministry of Education, Sri Lanka (MoE), Global School-Based Students Health Survey. Ministry of Health, Sri Lanka.,(Accessed 23/03/2022). https://extranet.who.int/ncdsmicrodata/index.php/catalog/30.2008

21. Ministry of Education, Sri Lanka (MoE). School Canteen Policy, Sri Lanka. Ministry of Education Sri lanka. 2006.

22. WHO, Measuring change in nutritional status. Annex 1: Standardization procedures for the collection of weight and height data in the field. Geneva: World Health Organization, 2008.. https://www.who.int/childgrowth/standards/Reliability_anthro.pdf?ua=1 1983. (Accessed 22/03/2022)

23. Liese A, Weis KE, Pluto D, Smith E, Lawson A. Food Store Types, Availability, and Cost of Foods in a Rural Environment. Journal of the Academy of Nutrition and Dietetics. 2007; 107: 1916–1923.

24. Rathi N, Riddle L, Worsley A. Parents’ and Teachers’ Views of Food Environments and Policies in Indian Private Secondary Schools. International Journal of Environmental Research and Public Health. 2017; 15: 1532.

25. Datar A, Nicosia N. Junk Food in Schools and Childhood Obesity. Journal of policy analysis and managemen. 2012; 31: 312–337.

26. Virtanen M, Kivimäki H, Ervasti J, Oksanen T, Pentti J, Kouvonen A. J. et al. Fast-food outlets and grocery stores near school and adolescents’ eating habits and overweight in Finland. European Journal of Public Health. 2015; 25.

27. Roudsari HA, Vedadhir A, Amiri P, Kalantari N, Omidvar N, Eini-zinab H, Hani SM. Psycho-Socio-Cultural Determinants of Food Choice: A Qualitative Study on Adults in Social and Cultural Context of Iran. Iranian journal of psychiatry. 2017; 12: 241–250.

28. Jayasinghe JM, Silva LP. Fast Food Consumption and Health Status of Students of a University in Sri Lanka. Journal of Food and Agriculture, 2014; 7: 38–50.

29. Skidmore ML, Welch AA, Sluijs EV, Jones PA, Griffin S, Cassidy A. Impact of neighbourhood food environment on food consumption in children aged 9-10 years in the UK SPEEDY (Sport, Physical Activity and Eating behaviour: Environmental Determinants in Young People) Study”. Public Health Nurition. 2010; 13: 1022–1030

30. Park S, Choi Y, Wang Y, Colantuoni E, Gittelsohn J. School and Neighborhood Nutrition Environment and Their Association With Students’ Nutrition Behaviors and Weight Status in Seoul, South Korea. Journal of Adelescent Health. 2013; 53: 655–662

31. Tang X, Ohri-vachaspati P, Abbott JK, Aggarwal R, Tulloch DL, Lloyd K, Yedidia M.J. Associations between food environment around schools and professionally measured weight status for middle and high school students.Childhood obesity. 2014; 10 : 511–517.

32. Mheroux M, Iannotti RJ, Curriec D, Pickett W, Janssen I.The food retail environment in school neighborhoods and its relation to lunchtime eating behaviors in youth from three countries. Health and Place. 2010; 18: 1240–1247.

33. Burgoine T, Jones AP, Namenek RJ, Benjamin SE. Associations between BMI and home, school and route environmental exposures estimated using GPS and GIS: do we see evidence of selective daily mobility bias in children?.Int J Health Geogr. 2015; 14.

34. García O, Tenorio Y, Ronquillo D, Ponce M. et al. Use of GIS to Measure Food Environment and Its Relationship with Obesity in School-aged Children in Mexico. Current Developments in Nutrition. 2019; P04–142-19.

35. Horst K, Timperio A, Crawford D, Roberts R, Brug J, Oenema A. The school food environment: associations with adolescent soft drink and snack consumption. American Journal of Preventive Medicine. 2008; 35(3), 217–223.

36. Buck C, Bornhorst C, Pohlabeln H, Huybrechts I, Pala V, Reisch L, Pigeot I. Clustering of unhealthy food around German schools and its influence on dietary behavior in school children: a pilot study. International Journal of Behavioral Nutrition and Physical Activity. 2013;10.

37. Ekanayake ULN. and Wijesinghe DGNG. Junk food consumption, physical activity and nutritional status of adolescent school children: A case study in Rathnapura District of Sri Lanka. Tropical Agricultural Research. 2021; 32(1): 105–113

38. Poti JM, Duffey KJ and Popkin BM. The association of fast food consumption with poor dietary outcomes and obesity among children: is it the fast food or the remainder of the diet?.Am J Clin Nutr. 2014; 99:162–171.

39. Tambalis K, Panagoitakos D, Psarra G, and Sidossis L. Association between fast-food consumption and lifestyle characteristics in Greek Children and Adolescents; Results from the EYZHN (National Action for Children’s Health) programme. Public Health Nutrition. 2018; 21(18): 3386–339).

40. Shier V, Nicosia N, Datar A. Neighborhood and home food environment and children’s diet and obesity: Evidence from military personnel’s installation assignment. Soc Sci Med. 2016 Jun;158:122–31.

41. Smagge BA, Velde LA, and Jong JC. The food environment sourrounding primary schools in a diverse urban area in Netherland: Linking fastfood density and proximity to neighborhood disadvantage and Childhood overweight prevalence. Front Public health. 2022; 10:

42. NDDCB, National Authority of Tobacco and Alcohol (NATA act no; 27 of 2006), (Accessed 14/02/2022). http://www.nddcb.gov.lk/Docs/acts/NATA%20Act%20English.pdf.

